# Physiological Effect of Prone Positioning in Mechanically Ventilated SARS- CoV-2 Infected Patients with Severe ARDS: Preliminary Analysis of an Observational Study

**DOI:** 10.1101/2020.09.16.20195958

**Authors:** Avishek Roy, Srikant Behera, Aparna Pande, Anirban Bhattacharjee, Amrita Bhattacharyya, Dalim K Baidya, Rahul K Anand, Bikash R Ray, Rajeshwari Subramaniam, Souvik Maitra

**Affiliations:** Department of Anaesthesiology, Pain Medicine & Critical Care, All India Institute of Medical Sciences, New Delhi

**Keywords:** SARS- CoV-2, ARDS, COVID-19, prone position, respiratory mechanics

## Abstract

Prone position ventilation has been shown to decrease mortality and improve oxygenation in ARDS patients. With best of our knowledge, no study reported physiological effect of prone position in SARS-CoV-2 infected ARDS patients. In this prospective observational study, data of n=20 consecutive laboratory confirmed SARS-CoV-2 patients with severe ARDS as per Berlin definition was included. Data of 20 patients analyzed with a median (Interquartile range, IQR) age of 56 (45.5-67) y and median (IQR) P/F ratio of 56 (54-66) with a median (IQR) PEEP of 12 (12-14) before initiation of prone position. Seventy-five percentage (95% CI 53.1-88.8) patients were prone responders at 16h prone session and 50 (95% CI 29.9-70.1) % patients were sustained responders. There was a significant decrease in plateau airway pressure (p<0.0001), peak airway pressure (p<0.0001) and driving pressure(p<0.0001) and increase in static compliance (p=0.001), P/F ratio (p<0.0001), PaO_2_ (p=0.0002)and SpO_2_ (p=0.0004) at 4h and 16h since initiation of prone session and also after return of supine position. Prone position in SARS-CoV-2 infected severe ARDS patients is associated with improvement in lung compliance and oxygenation in two-third of the patients and persisted in half of the patients.

## Introduction

Since the diagnosis of first case in December 2019, SARS-CoV-2 has infected more that 16 million and caused fatality in more than 600,000 people worldwide. Early data from China reported that around 5% of all laboratory confirmed cases become critically ill. [1]. Although the overall case fatality rate (CFR) ranges between 2.3 to 7.4 %, mortality in critically ill can reach 26% [2].

Early reports from Italy indicated that among the different phenotypes of ARDS in SARS-CoV-2 pneumonia that have been proposed, the ‘L’ phenotype has normal lung compliance and lung weight but leads to hypoxemia due to the loss of hypoxic pulmonary vasoconstriction, later it progresses to ‘H’ phenotype with low compliance and increased lung weight [3].

Prone position ventilation has been shown to decrease mortality and improve oxygenation in ARDS patients. In mechanically ventilated patients of severe ARDS with PaO_2_/FiO_2_ (P/F) ratio < 150, prone position for at least 4 days decreased 28-day mortality by almost 50% [4]. Prone position reduces lung strain and stress, leads to more homogenized distribution of lung aeration and recruitment of dorsal alveoli, thus, leading to improvement in oxygenation [5].

With best of our knowledge, no study reported physiological effect of prone position in SARS-CoV-2 infected ARDS patients. Hence, in this preliminary analysis of an observational study, physiological effect of prone position in SARS-CoV-2 infected severe ARDS patients have been reported.

## Methods

Permission from the Institute Ethics Committee was obtained before recruitment of first patient and consent was obtained from legally acceptable representative of all recruited patients. In this prospective observational study, data of n=20 consecutive laboratory confirmed SARS-CoV-2 patients with severe ARDS as per Berlin definition was included.

As per ICU protocol, in the absence of contraindication, all mechanically ventilated ARDS patients with PaO_2_/ FiO_2_< 150 were placed in at least 16h/day prone position for consecutive days till the criteria is met. Demographic characteristics, baseline respiratory mechanics and blood gas data were collected before initiation of prone position, after 4h and 16h of prone position and after 4h of return of supine position. Positive end expiratory pressure (PEEP) was titrated as per *ARDSNet protocol* PEEP-FiO_2_ table. Prone responders were defined by 20% increase in PaO_2_/ FiO_2_ ratio during the prone session and sustained responders were defined by 20% increase in PaO_2_/ FiO_2_ ratio 4h after return of supine position.

All collected data were entered in a Microsoft Excel datasheet. Categorical data were presented as as absolute numbers or percentages and non-parametric data were presented as median and IQR (inter-quartile range). Unrelated data (between prone responders and non-responders) were compared by Mann-Whitney U test or Chi-square test as applicable. Longitudinal variables were compared by Friedman’s test and multiple comparisons were performed by Dunn’s test. A two-sided p value < 0.05 was considered as significant. All statistical analyses were performed using GraphPad Prism version 8.0.0 for Mac OS, (GraphPad Software, San Diego, California USA).

## Results

In this observational study, data of 20 patients analyzed with a median (IQR) age of 56 (45.5-67) y and median (IQR) predicted body weight of 60 (55-62.5) kg. Baseline respiratory mechanics data is presented in table 1. All included patients had severe ARDS with median (IQR) P/F ratio of 56 (54-66) with a median (IQR) PEEP of 12 (12-14) before initiation of prone position. Median (IQR) SOFA score was 7.5 (5.5-9) at the time of inclusion. Seventy-five percentage (95% CI 53.1-88.8) patients were prone responders at 16h prone session and 50 (95% CI 29.9-70.1) % patients were sustained prone responders after return to supine position. Prone responders had significantly higher baseline respiratory compliance (p=0.03,Mann Whitney U test) but all other respiratory and blood gas variables were similar between responders and non-responders.

**Table 1:**
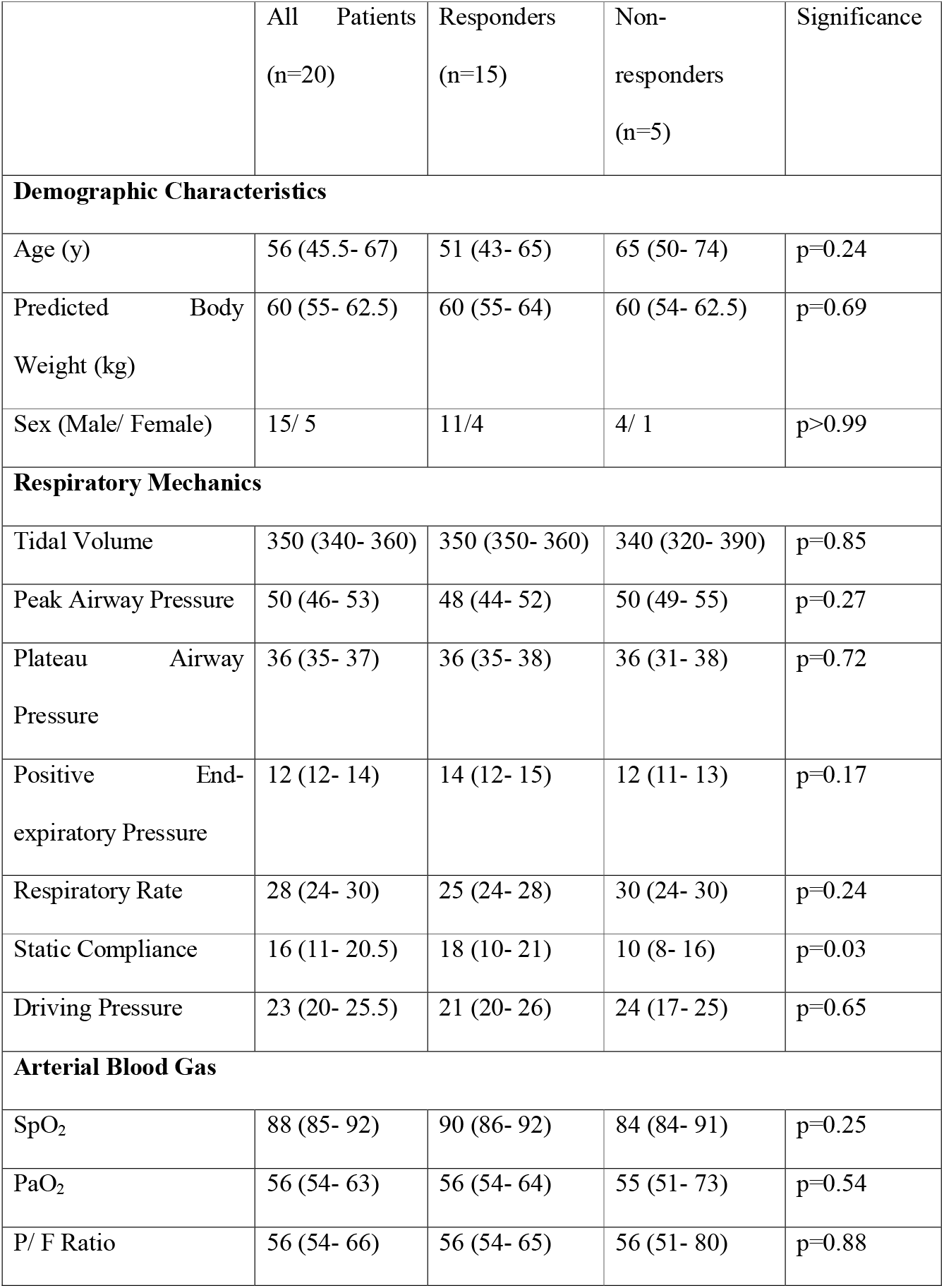
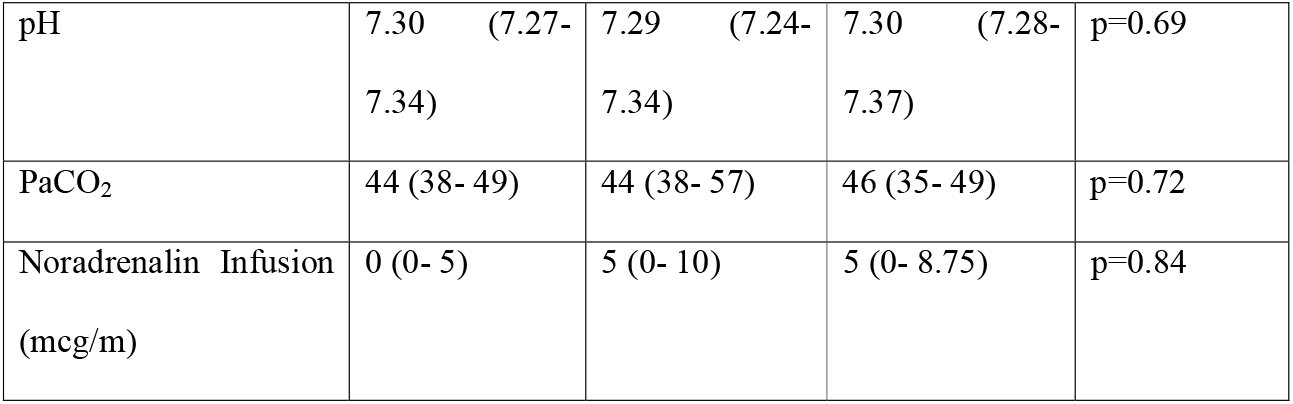
Respiratory Mechanics data at the time of enrolment (before prone position)

There was a significant decrease in plateau airway pressure (p<0.0001), peak airway pressure (p<0.0001) and driving pressure(p<0.0001) and increase in static compliance (p=0.001) at 4h and 16h since initiation of prone session and also after return of supine position. Change in respiratory mechanics parameters from baseline are reported in figure 1. P/F ratio (p<0.0001), PaO_2_ (p=0.0002) and SpO_2_ (p=0.0004) increased from baseline and persisted in supine position also (figure 1). Noradrenalin requirement didn’t change during the prone session (p=0.20). Percentages of changes in static compliance significantly correlated with P/F ratio after return of supine position (r^2^=0.62, p=0.0034) but not at 4h (p=0.14) and 16h (p=0.20). Percentages of changes in P/F ratio and driving pressure at 16h (r^2^=-0.47, p=0.04) and after return of supine position (r^2^=-0.59, p=0.0089) were significantly correlated; but no correlation was found at 4h (p=0.09).

**Figure 1:**
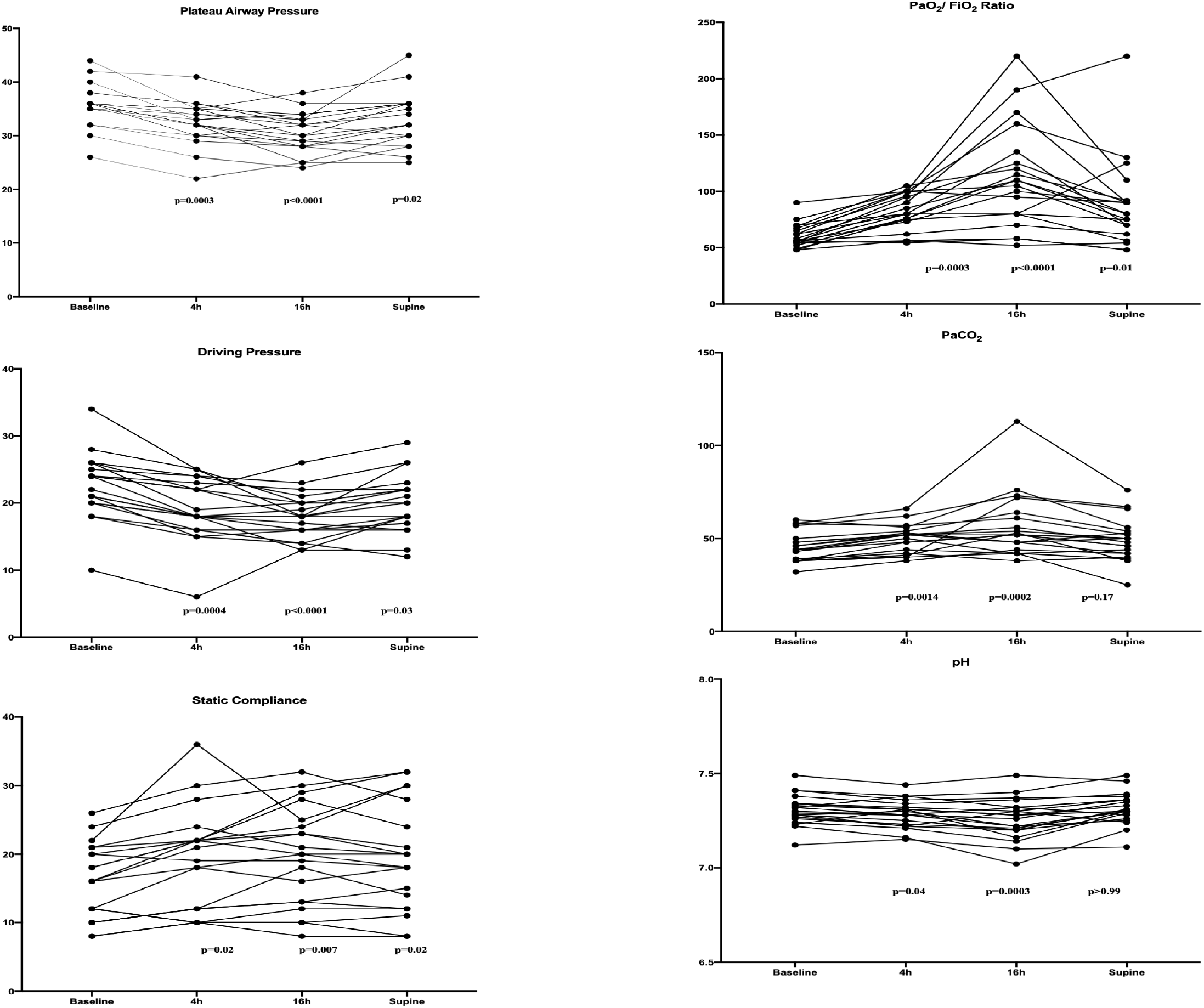
Before-after line graph showing respiratory mechanics and blood gas parameters in all patients [p values for Dunn’s multiple comparison (baseline vs other time points) after Friedman’s test reported]

Baseline static compliance was a predictor of prone response with reasonable accuracy [AUROC (95% CI) 0.82 (0.59-1.00)]. Static compliance < 14 predicted no response from prone position with sensitivity (95% CI) and specificity (95% CI) of 80 (37.6-99) % and 73.3 (48.1-89.1) % respectively. At the time of writing of this manuscript, 4 patients [proportion (95% CI) 22.2 (9-45.2) %] were discharged from hospital and 2 patients were still in the hospital.

## Discussion

We have found that around 75% of the SARS-CoV-2 infected patients with severe ARDS responded with 16h prone position in terms of oxygenation. Overall there is improvement in lung mechanics in terms of static compliance, driving pressure and plateau pressure without any changes in the hemodynamic support. In our series, all the included patients had ‘stiff lung’ as evident by low static compliance. Previous studies reported a variable change in respiratory system compliance in prone position in ARDS patients [6], whereas we have found a significant decrease in driving pressure and static compliance. Recruitment of the dorsal lung region was the biologically plausible mechanism of improvement in static compliance as both driving pressure and compliance were correlated with change in P/F ratio [7]. We have found that these correlations were present even after return of supine position which indicated a sustained lung recruitment in SARS-CoV-2 infected patients. Determination of baseline static compliance is important as it was a predictor of absence of response from prone position and these patients may be subjected to extra-corporeal membrane oxygenation early in the course of disease. Our study has few limitations such as sample size was small, and we couldn’t assess the effect of prone position on chest wall and lung compliance separately as esophageal manometry was not used.

## Conclusion

Prone position in SARS-CoV-2 infected severe ARDS patients is associated with improvement in lung compliance and oxygenation in two-third of the patients and persisted in half of the patients.

## Data Availability

Anonymous data will be available from the corresponding author on reasonable request.

## References

1. Wu Z, McGoogan JM. Characteristics of and important lessons from the coronavirus disease 2019 (COVID-19) outbreak in China: summary of a report of 72 314 cases from the Chinese Center for Disease Control and Prevention. Jama. 2020 Apr 7;323:1239–42.

2. Yang X, Yu Y, Xu J, Shu H, Liu H, Wu Y, Zhang L, Yu Z, Fang M, Yu T, Wang Y. Clinical course and outcomes of critically ill patients with SARS-CoV-2 pneumonia in Wuhan, China: a single-centered, retrospective, observational study. The Lancet Respiratory Medicine. 2020 Feb 24.

3. Grasselli G, Zangrillo A, Zanella A, Antonelli M, Cabrini L, Castelli A, Cereda D, Coluccello A, Foti G, Fumagalli R, Iotti G. Baseline characteristics and outcomes of 1591 patients infected with SARS-CoV-2 admitted to ICUs of the Lombardy Region, Italy. JAMA. 2020;323:1574–81.

4. Guérin C, Reignier J, Richard JC, Beuret P, Gacouin A, Boulain T, Mercier E, Badet M, Mercat A, Baudin O, Clavel M. Prone positioning in severe acute respiratory distress syndrome. New England Journal of Medicine. 2013;368:2159–68.

5. Mentzelopoulos Mentzelopoulos SD, Roussos C, Zakynthinos SG. Prone position reduces lung stress and strain in severe acute respiratory distress syndrome. European Respiratory Journal. 2005;25:534–44.

6. Kallet RH. A Comprehensive Review of Prone Position in ARDS. Respir Care. 2015;60:1660–1687.

7. Lee DL, Chiang HT, Lin SL, Ger LP, Kun MH, Huang YC. Proneposition ventilation induces sustained improvement in oxygenation in patients with acute respiratory distress syndrome who have a large shunt. Crit Care Med 2002;30:1446–1452.

